# Association of COVID-19 vaccination with risks of hospitalization due to cardiovascular and other diseases: A study of the UK

**DOI:** 10.1101/2021.08.15.21262097

**Authors:** Yong Xiang, Yaning Feng, Jinghong Qiu, Ruoyu Zhang, Hon-Cheong So

## Abstract

**Background:** Vaccines for COVID-19 represent a major breakthrough. However, worries about adverse effects led to vaccine hesitancy in some people. On the other hand, as COVID-19 may be associated with various sequelae, vaccination may protect against such sequelae via prevention of infections and severe disease.

**Methods:** We leveraged the UK-Biobank (UKBB) and studied associations of at least one dose of COVID-19 vaccination (BioNTech-BNT162b2 or Oxford-AstraZeneca-ChAdOx1) with short-term hospitalizations from cardiovascular and other selected diseases (*N*=393,544; median follow-up*=*54 days among vaccinated). Multivariable Cox and Poisson regression analyses were performed. We also performed adjustment using prescription-time distribution matching (PTDM) and prior-event rate ratio (PERR). PERR minimizes unmeasured confounding by comparing event hazards before introduction of vaccination.

**Results:** We observed that COVID-19 vaccination(at least one dose), when compared to no vaccination, was associated with reduced short-term risks of hospitalizations from stroke(hazard ratio[HR]=0.178, 95% CI: 0.127-0.250, *P=*1.50e-23), venous thromboembolism (VTE) (HR=0.426, CI: 0.270-0.673, *P=*2.51e-4), dementia(HR=0.114, CI: 0.060-0.216; *P=*2.24e-11), non-COVID-19 pneumonia(NCP) (HR=0.108, CI: 0.080-0.145; *P=*2.20e-49), coronary artery disease (CAD) (HR=0.563, CI: 0.416-0.762; *P=*2.05e-4), chronic obstructive pulmonary disease (COPD) (HR=0.212, CI: 0.126-0.357; *P=*4.92e-9), type-2 diabetes (T2DM) (HR=0.216, CI: 0.096-0.486, *P=*2.12e-4), heart failure (HR=0.174, CI: 0.118-0.256, *P=*1.34e-18) and renal failure (HR=0.415, CI: 0.255-0.677, *P=*4.19e-4), based on Cox regression models. Among the above results, reduced hospitalizations for stroke, heart failure, NCP and dementia were consistently observed across all analyses, including regression/PTDM/PERR.

**Conclusions:** Taken together, this study provides further support to the safety and benefits of COVID-19 vaccination, and such benefits may extend beyond reduction of infection risk or severity per se. However, causal relationships cannot be concluded and further studies are required to verify the findings.

## Introduction

More than 550 million confirmed cases of COVID-19 and >6.3 million fatalities have been reported as at 13-July-2022(https://coronavirus.jhu.edu/map.html). Vaccines for COVID-19 have been developed at an unprecedented speed, and offer hope to reduce the burden of this pandemic. Nevertheless, vaccine hesitancy is a major hurdle, and some may worry about adverse effects or exacerbation of existing diseases^1,2^. There were case reports of fatalities after vaccination when COVID-19 vaccines were first introduced^3,4^, which has led to concerns about the safety of vaccination among some individuals. However, so far there has been no direct evidence that COVID-19 vaccination is causally linked to increased risks of mortality in general.

On the other hand, as COVID-19 may be associated with various complications such as cardiovascular events, thromboembolism, renal failure etc.^5,6^, it is reasonable to hypothesize that COVID-19 vaccination may reduce the risks of these complications. Notably, it has been shown in many reports and meta-analysis that influenza vaccination is associated with reduced cardiovascular risks and mortality^7-10^. Reduction of flu infection is believed as the major mechanism; however, flu vaccines may also promote plaque stabilization^11,12^ and nitric oxide production^13^. It is possible that COVID-19 vaccination may also reduce cardiovascular risks and risks from other relevant sequelae.

Here we investigate the association of COVID-19 vaccination with hospitalization from cardiovascular and other diseases. Cardiovascular diseases(CVD) are chosen as they are leading causes of mortalities worldwide, and the protective effects of flu vaccines against CVD lead us to hypothesize similar effects for COVID-19 vaccines. We also included several other diseases likely linked to COVID-19 as complications/sequelae. For example, renal dysfunction is a common complication^14^ and is closely related to CVD. COPD exacerbation is common after viral infections^15^ and a relevant complication. Venous thromboembolism(VTE) is another known complication^16^. In addition, both the infection and vaccination involve interplay with the immune system, and therefore autoimmune diseases may be associated with infection and/or vaccination^17^. As for neurological sequelae, a recent study reported that across all neuropsychiatric disorders, risk of mortality from dementia was particularly elevated^18^. Here the association of COVID-19 vaccination with hospitalization from the above diseases will be investigated.

## Methods

### UK Biobank sample

The UK Biobank(UKBB) is a large-scale prospective cohort comprising ∼500,000 subjects aged 40–69 years when recruited in 2006–2010. The current age of subjects ranged from 50-87 years. For details of UKBB please see^19^. We included subjects with available General Practice(GP) records(which covers vaccination records) under the TPP and EMIS systems(sample size *N*=393,544). This analysis was conducted under project number 28732.

### Outcome definition

Hospitalization records were extracted from the Hospital Episode Statistics(HES) of UKBB. Detailed descriptions of the HES can be found in https://biobank.ndph.ox.ac.uk/ukb/ukb/docs/HospitalEpisodeStatistics.pdf. The inpatient core and diagnoses datasets were updated to 31-Mar-2021. The diagnosis codes and corresponding dates were summarized based on each participant’s inpatient record. All diagnosis codes were converted to 3-character ICD-10 codes. We followed the mapping strategy as described in https://biobank.ndph.ox.ac.uk/showcase/showcase/docs/first_occurrences_outcomes.pdf.

The ICD-codes for defining each disease outcome were listed in Table_S1. The disease outcomes(hospitalization) studied included coronary artery disease(CAD), atrial fibrillation(AF), heart failure(HF), hypertension(HTN), stroke, renal failure(RF), type 2 diabetes mellitus(T2DM), venous thromboembolism(VTE), systemic and organ-specific autoimmune diseases, chronic obstructive pulmonary disease(COPD), non-COVID-19 pneumonia(NCP) and dementia.

Only primary causes of admission were considered. Diagnosis given before the date of first vaccination were regarded as medical history of comorbidity; diagnoses given afterwards were treated as new hospitalizations. For better statistical power, we primarily present the results from any hospitalizations from the diseases. However, we also conducted further separate analysis on new-onset(no prior history) and recurrent/relapsed diseases(known history of the disease) as outcomes.

### Covariates

The full set of covariates(Table_S2) included basic demographic variables(age, sex, ethnic group), comorbidities(CAD, stroke, T2DM, HTN, AF, COPD, dementia, history of cancer, chronic kidney disease[CKD], history of pneumonia), risk factors of cardiometabolic disorders(lipids/glucose/HbA1c), disorders of the immune system(autoimmune diseases/immunodeficiencies/drug history of immunosuppressants), general health indicators(number of medications prescribed by GP and number of hospitalizations in the past year, number of non-cancer illnesses), anthropometric/obesity measures(body mass index[BMI]/waist-circumference), socioeconomic status(Townsend Deprivation index) and lifestyle risk factor(smoking). For disease traits, we included information from primary care data, hospital inpatient data, ICD-10 diagnoses(code-41270) and self-reported illnesses(code-20002) and incorporated data from all waves of follow-ups. The strategy of integrating all the diagnosis records were based on information provided by UKBB(https://biobank.ndph.ox.ac.uk/showcase/showcase/docs/first_occurrences_outcomes.pdf). Subjects with no records of a disease from any data source were regarded as having no history of the disease.

We note that the number of events may be small for some diseases, yet the number of covariates is relatively large. In view of this, we performed regression analyses using two strategies:(1)’Basic’ model: it includes age, sex, prior infection, number of hospitalizations and medications received in the past year as covariates;(2)’advanced’ model: under this strategy, all covariates were considered. We first included the covariates under the basic model. For the rest of the covariates, we first performed univariate screening with each covariate; those with(nominally) significant associations(p<0.05) with hospitalization were selected into the final model. This approach followed ref^20^, which was shown by theory and simulations to control the proportion of false-positive predictors.

### Missing covariate data

Missing values were imputed with R-package ‘missRanger’. The program is based on missForest, an iterative imputation approach based on random forest. It is widely used and was shown to produce low imputation errors and good performance in predictive models^21^. The missing rate of each covariate and the OOB(out-of-bag) error of imputation are listed in Table_S3.

### COVID-19 infection status

Information regarding COVID-19 data in the UKBB was given at https://biobank.ndph.ox.ac.uk/showcase/exinfo.cgi?src=COVID19. Briefly, the latest COVID test results were downloaded from UKBB(last update 21-Jul-2021). The strategy of identifying the positive of SARS-CoV-2 can be found in (https://biobank.ndph.ox.ac.uk/showcase/showcase/docs/casecontrol_covidimaging.pdf). Untested subjects or subjects without any positive results were treated as uninfected.

### Vaccination status definition

Vaccination status was extracted from the TPP and EMIS General Practice clinical records(TPP last update 21 Jul 2021; EMIS last update 10 Aug 2021). During the period of study, all subjects received either the BioNTech BNT162b2 or Oxford-AstraZeneca ChAdOx1 nCoV-19 vaccine. Please also refer to Figure 1 for an overview of our analysis workflow.

**Figure 1.**
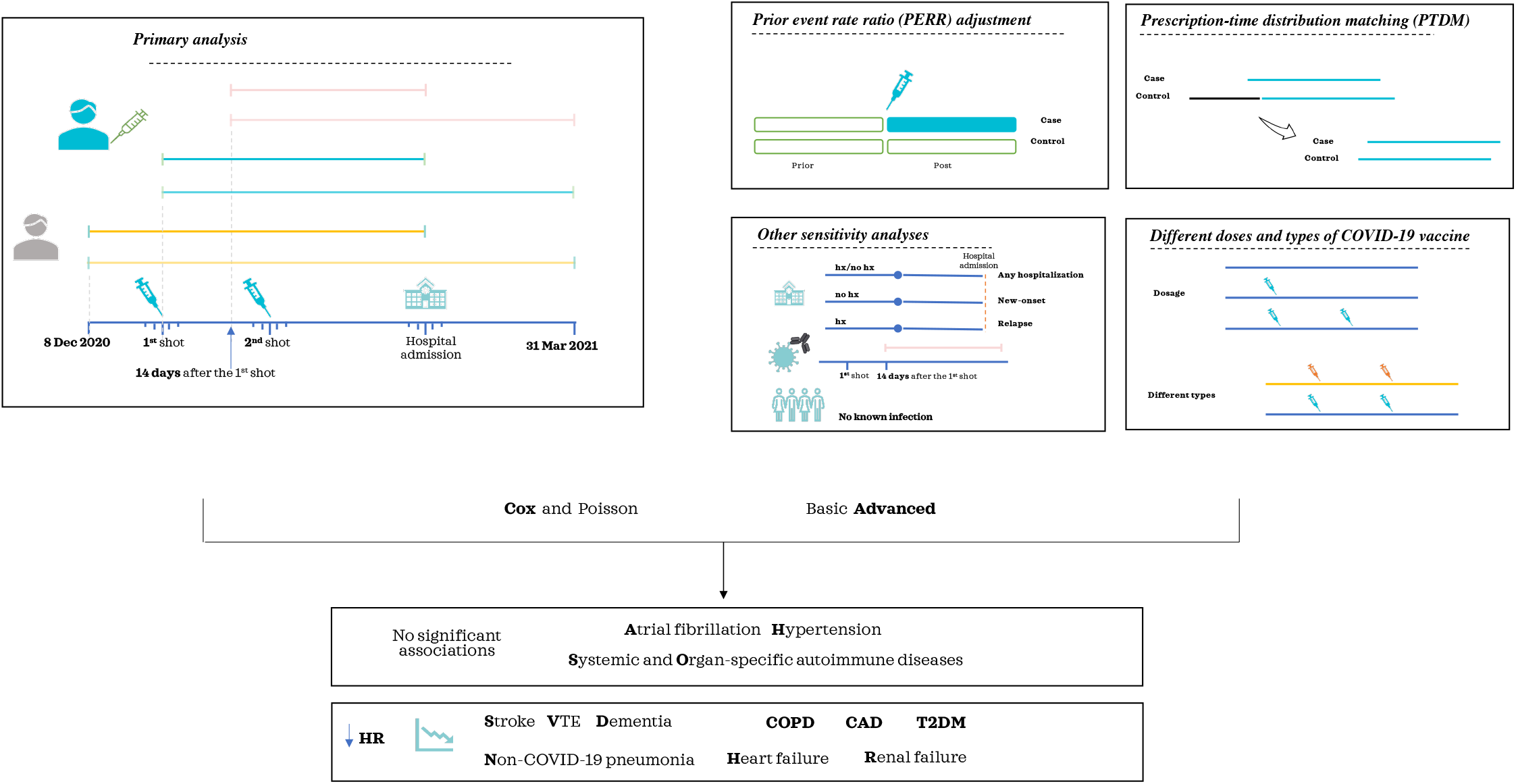
An overview of the analytic workflow. For the primary analysis, we mainly investigated the association of as least one dose of COVID-19 vaccine with hospitalization from various diseases. For sensitivity analyses, we conducted prior event rate ratio (PERR) adjustment, prescription-time distribution matching (PTDM) adjustment, separate analyses for different dosages and types of COVID-19 vaccines and other sensitivity analyses as described in the text. Cox and Poisson regression model were employed for association testing. We also considered both ‘basic’ and ‘advanced’ modeling; the former model contains a few basic covariates while the latter considers a larger number of covariates. Please refer to the main text for details.

### Statistical analysis

For details please also refer to Supplementary Text.

Briefly, Cox and Poisson regression were performed. Time-to-hospitalization was treated as outcome in Cox regression, controlling for other covariates. We checked the proportional hazards(PH) assumption by R function cox.zph. For Poisson regression, presence/absence of event was considered as outcome, with ‘offset’ specified(=number of days of follow-up) to account for differences in follow-up durations. For vaccinated subjects, the start-date was date-of-first-vaccination; for unvaccinated subjects, the start-date was set at 8-Dec-2020, the date COVID-19 vaccines were first deployed in the UK. The end of follow-up was set at 31-Mar-2021, as UKBB records of hospitalization were updated up to that date at the time of analysis. All statistical analysis were performed by R(3.6.1).

#### Prior event rate ratio(PERR) adjustment

PERR adjustment is a methodology to minimize the effect of unmeasured confounders^22-24^. The basic idea is to adjust for the differences in event rates between the two(treated and untreated) groups *before* treatment initiation, such that baseline differences between the two groups are accounted for.

#### Prescription-time distribution matching(PTDM)

Individuals who die shortly after start-date will not have the opportunity to receive vaccination, and this can result in a survival advantage(bias) for the treated(vaccinated) subjects. This bias is most substantial if mortality is considered as the outcome. Here we focused on hospitalization, so the effect of survival bias is less severe. A person can still get vaccinated after being hospitalized; however, hospitalization is a risk factor of mortality, hence some degree of survival bias may remain. Here we employed the PTDM approach to reduce survival bias^25^. Briefly, for each unvaccinated person, the start-date(time0) is randomly chosen from the start-dates of the vaccinated group to ensure a similar distribution.

#### Analyses accounting for number of doses received and types of COVID-19 vaccine

Further stratification analyses were conducted to determine whether the effectiveness of the COVID-19 vaccine varied depending on the dose and type of vaccine. We compared two-dose vaccination(N=43,659 and the date of receiving the second dose was treated as its ‘start-date’) vs no vaccination(*N*=22,482), one dose only(*N*=327,403) vs no vaccination, and the effect of two doses vs only one dose. The number of doses received were counted as at 31-Mar-2021. Despite that a proportion of subjects did not have details of the vaccine type, we still identified that a total of 87,778 subjects received the BioNTech BNT162b2 vaccine and 143,576 individuals received the Oxford-AstraZeneca ChAdOx1 nCoV-19 vaccines. Comparisons were made between different types of COVID-19 vaccine against those with no vaccination, as well as between the two types of vaccines.

#### Other analysis to check for robustness of findings

We performed various analyses to verify the robustness findings under different modeling strategies/assumptions:(1)While we primarily focused on all hospitalizations, we also conduct stratified analysis for new-onset and recurrent diseases, as described above;(2)for counting the ‘start-date’, we primarily consider it as the date of vaccination, as it is possible that some side-effects can occur early. However, we also performed another set of analysis with start-date set at 14 days after the 1^st^ vaccination, since the protective effects of vaccines may only be apparent ∼2 weeks later;(3)we also repeated the analyses limited to those with no recorded history of infection all along. Note that as UKBB subjects were not routinely screened for infection, a substantial proportion of asymptomatic, mild or moderate infections may not be captured. Analysis(3) may reveal whether vaccination may confer protective effects against other diseases presumably via protection against milder infections.

## Results

We will primarily present our findings with at least one dose of vaccination, and results from Cox regression with start-date defined as the day of(first) vaccination. In general, we found results from Cox or Poisson regression, or different start-dates to be similar. We will also present the results with PERR and PTDM adjustment. If not further specified, we will describe the results under the ‘advanced’ model(which considers full set of covariates), although the results from basic and advanced models were generally similar. The main findings are mostly robust to different modeling strategies. The full results of all analyses(including sensitivity analysis) are presented in Tables_S4-S9 and Figures_S1-S8. The median length of follow-up for the vaccinated group was 54 days, while the corresponding length was 113 days in the unvaccinated group without PTDM adjustment and 55 days with PTDM.

### Association of hospitalization with COVID-19 vaccination status(at least one dose of vaccine)

The main results are listed in Table_1 and full results in Tables_S4-S5. Please also refer to Figure_2. Here we first present the results based on Cox regression model(‘advanced’ model with consideration of all covariates and pre-screening), without further adjustments by PTDM or PERR. Considering all hospitalizations(regardless of history of the studied disease), COVID-19 vaccination was significantly associated with reduced hazards of hospitalization due to stroke(hazard ratio[HR]=0.178,CI:0.127-0.250,p=1.50e-23), VTE(HR=0.426,CI:0.270-0.673,p=2.51e-4), dementia(HR=0.114,CI:0.060-0.216;p=2.24e-11), non-COVID-19 pneumonia(NCP)(HR=0.108,CI:0.080-0.145;p=2.20e-49), CAD(HR=0.563,CI:0.416-0.762;p=2.05e-4), COPD(HR=0.212,CI:0.126-0.357;p=4.92e-9), T2DM(HR=0.216,CI:0.096-0.486,p=2.12e-4), heart failure(HR=0.174,CI:0.118-0.256,p=1.34e-18) and renal failure(HR=0.415,CI:0.255-0.677,p=4.19e-4). The results from the ‘basic’ regression model(with less covariates included) were similar, with slightly lower HR estimates(i.e. stronger protective associations)(Table_S7).

**Table 1.** Association of (at least one dose of) COVID-19 vaccination with hospitalization from various disorders

| Outcome | Coef | Exp(coef)<br>[HR] | SE(coef) | Z | P | LCI | UCI | No of<br>events in<br>unvac | Rate of events<br>in unvac<br>(per 100,000<br>person-days) | No of<br>events<br>in vac | Rate of<br>events<br>in vac<br>(per<br>100,000<br>person-days) |
| --- | --- | --- | --- | --- | --- | --- | --- | --- | --- | --- | --- |
| AF | -0.151 | 0.860 | 0.236 | -0.642 | 5.21E-01 | 0.542 | 1.364 | 23 | 0.93 | 274 | 1.44 |
| CAD | -0.574 | 0.563 | 0.155 | -3.713 | **2.05E-04** | 0.416 | 0.762 | 61 | 2.48 | 443 | 2.33 |
| COPD | -1.551 | 0.212 | 0.265 | -5.85 | **4.92E-09** | 0.126 | 0.357 | 22 | 0.89 | 63 | 0.33 |
| Dementia | -2.171 | 0.114 | 0.324 | -6.69 | **2.24E-11** | 0.06 | 0.216 | 18 | 0.73 | 28 | 0.15 |
| Heart Failure | -1.751 | 0.174 | 0.199 | -8.803 | **1.34E-18** | 0.118 | 0.256 | 41 | 1.66 | 109 | 0.57 |
| Non covid pneumonia | -2.226 | 0.108 | 0.151 | -14.773 | **2.20E-49** | 0.08 | 0.145 | 91 | 3.70 | 139 | 0.73 |
| Organ specific<br>autoimmune | 0.317 | 1.372 | 0.37 | 0.856 | 3.92E-01 | 0.665 | 2.833 | 9 | 0.37 | 125 | 0.66 |
| Renal Failure | -0.879 | 0.415 | 0.249 | -3.528 | **4.19E-04** | 0.255 | 0.677 | 28 | 1.14 | 112 | 0.59 |
| Stroke | -1.724 | 0.178 | 0.172 | -10.001 | **1.50E-23** | 0.127 | 0.25 | 54 | 2.19 | 173 | 0.91 |
| Systemic autoimmune | -0.362 | 0.696 | 0.51 | -0.709 | 4.78E-01 | 0.256 | 1.892 | 5 | 0.20 | 44 | 0.23 |
| T2DM | -1.533 | 0.216 | 0.414 | -3.704 | **2.12E-04** | 0.096 | 0.486 | 9 | 0.37 | 27 | 0.14 |
| VTE | -0.853 | 0.426 | 0.233 | -3.661 | **2.51E-04** | 0.27 | 0.673 | 30 | 1.22 | 135 | 0.71 |

Coef, coefficient from Cox regression; SE, standard error; HR, hazard ratio; LCI and UCI, lower and upper 95% confidence interval of HR. AF, atrial fibrillation; CAD, coronary artery disease; COPD, chronic obstructive pulmonary disease; HTN, hypertension; T2DM, type 2 diabetes; VTE, venous thromboembolism; vac, vaccinated group; unvac, unvaccinated group.
Results with $p < 0.05$ are in bold.
The results above were based on the 'advanced model' (i.e. consideration of all covariates with pre-screening). No further PERR or PTDM adjustments were made and for the vaccinated group day0 was defined as the date of vaccination.

**Figure 2.**
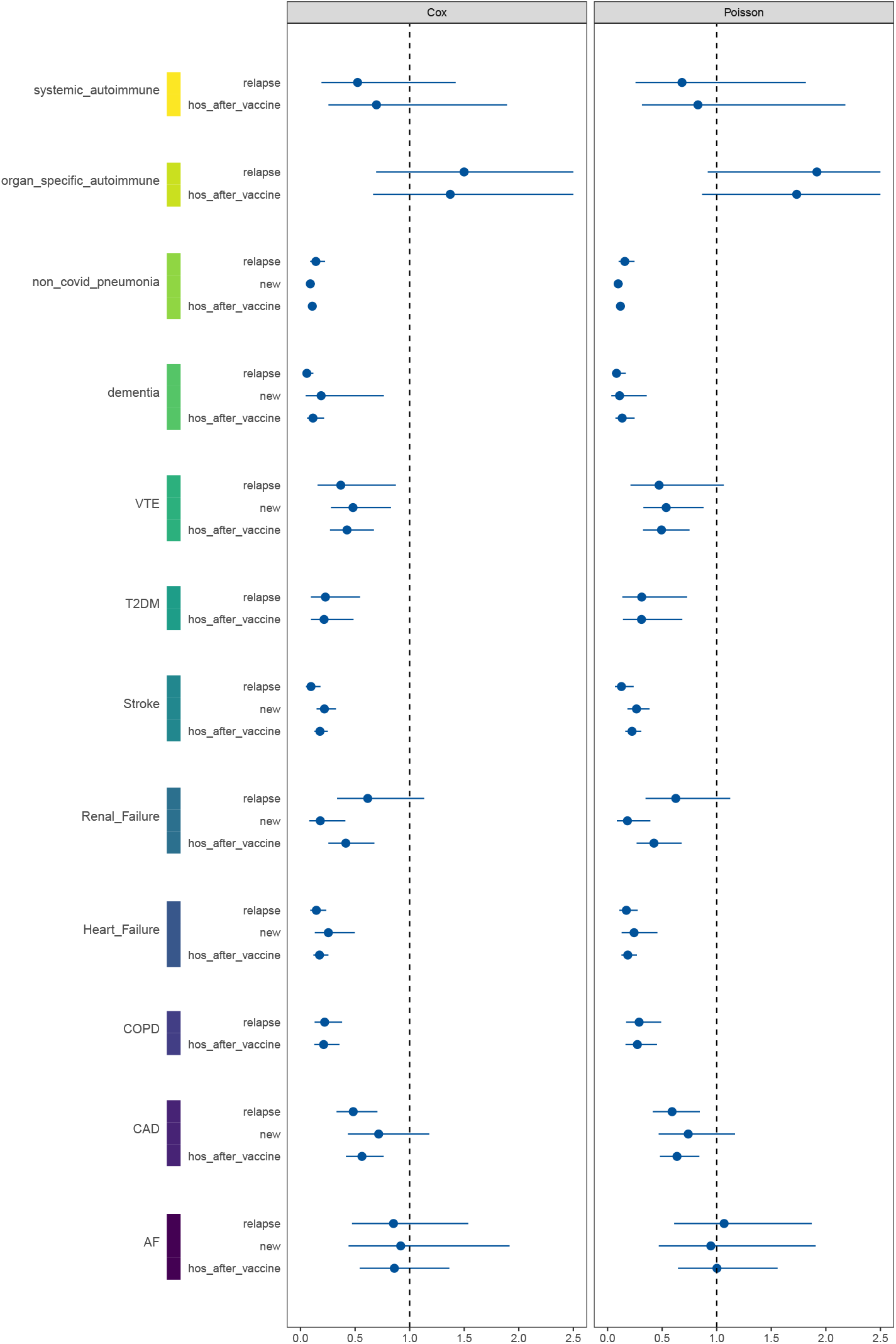
An overview of association of (at least one dose of) COVID-19 vaccination with hospitalization from various diseases. The analysis was conducted under the ‘advanced model’ with consideration of all covariates and pre-screening, without PTDM or PERR adjustments. Totally three types of outcomes were defined: hos_after_vaccination, all hospitalizations after vaccination; new-onset, hospitalizations from new-onset diseases (without prior history of the disease); relapse, hospitalizations among those with known history of the studied disease. To avoid unstable estimates from very few events, we present the results only if the number of events>=5 in both vaccinated and un-vaccinated groups. To preserve readability of the graph, the upper 95% confidence intervals of HR are truncated at 2.5.

Restricting the outcome to hospitalizations due to *new-onset* diseases(i.e. without prior history), we observed that vaccination was associated with reduced hazards of hospitalizations from stroke(HR=0.219,CI:0.147-0.325;p=8.40e-3), renal failure(HR=0.181,CI:0.080-0.411;p=4.36e-5), heart failure(HR=0.255,CI:0.131-0.498;p=6.08e-5), VTE(HR=0.481,CI:0.279-0.829;p=8.40e-3), dementia(HR=0.189,CI:0.047-0.764;p=1.94e-2) and NCP(HR=0.090,CI:0.061-0.131;p=9.88e-36). Note that the number of events was too small for further analyses for some diseases. There were only 5 and 8 new hospitalizations due to dementia in the unvaccinated and vaccinated groups, so the results should be viewed with caution.

On the other hand, if we consider hospitalizations due to *recurrent* disease(i.e. those with known history of the disease) as outcome, vaccination was associated with decreased hospitalization hazards due to stroke(HR=0.096,CI:0.050-0.183;p=2.04e-3), CAD(HR=0.483,CI:0.330-0.705;p=1.68e-4), COPD(HR=0.221,CI:0.129-0.381;p=5.40e-8), heart failure(HR=0.145,CI:0.089-0.236;p=9.82e-15), NCP(HR=0.141,CI:0.088-0.225), VTE(HR=0.369,CI:0.156-0.874, p2.35e-2) and dementia(HR=0.058,CI:0.028-0.118;p=5.82e-15). In general, the protective associations were more prominent for recurrent cases when compared to all hospitalizations without consideration of disease history.

The results from the ‘basic’ regression models for hospitalizations due to new or recurrent disease were similar, with slightly lower HR estimates(Table_S7). The same set of significant associations were observed. For completeness, we also present the results when all covariates were included without pre-screening(Table_S8). We show by default only the results without warnings of convergence issues from the “coxph” function.

We also repeated the analysis using Poisson regression which models the incidence rate of hospitalization. The results were highly similar to those from Cox regression, with similar significant associations observed(Tables_S4-5).

### With PERR adjustment to reduce residual confounding

The primary results with PERR adjustment(number of bootstraps=1000; the prior-to-vaccination period restricted to 1-Jan-2019 to 7-Dec-2020) are listed in Table_2 andTable_S6. Please also refer to Figure 3 and Figure S1. After PERR adjustment, COVID-19 vaccination was significantly associated with reduced hazards of hospitalization due to CAD(HR=0.517,CI:0.327-0.817;p=4.70e-03), COPD(HR=0.317,CI:0.138-0.726;p=6.60e-03), dementia(HR=0.406,CI:0.180-0.919;p=3.06e-02), heart failure(HR=0.305,CI:0.179-0.519;p=1.18e-05), NCP(HR=0.191,CI:0.123-0.295;p=1.02e-13) and stroke(HR=0.222,CI:0.130-0.382;p=4.94e-08).

**Table 2.** Association of (at least one dose of) COVID-19 vaccination with hospitalization from various disorders after PERR (prior event rate ratio) adjustment

| Outcome | beta-post | beta-pre | beta-diff | boot-se | HR | LCI | UCI | Z | P |
| --- | --- | --- | --- | --- | --- | --- | --- | --- | --- |
| AF | -0.151 | 0.199 | -0.35 | 0.288 | 0.704 | 0.401 | 1.239 | -1.217 | 2.24E-01 |
| CAD | -0.574 | 0.085 | -0.66 | 0.233 | 0.517 | 0.327 | 0.817 | -2.827 | **4.70E-03** |
| COPD | -1.551 | -0.401 | -1.15 | 0.423 | 0.317 | 0.138 | 0.726 | -2.716 | **6.60E-03** |
| Dementia | -2.171 | -1.27 | -0.9 | 0.416 | 0.406 | 0.18 | 0.919 | -2.162 | **3.06E-02** |
| Heart Failure | -1.751 | -0.563 | -1.188 | 0.271 | 0.305 | 0.179 | 0.519 | -4.382 | **1.18E-05** |
| HTN | 0.363 | 0.103 | 0.261 | 3.158 | 1.298 | 0.003 | 633.389 | 0.083 | 9.34E-01 |
| Non covid pneumonia | -2.226 | -0.57 | -1.656 | 0.223 | 0.191 | 0.123 | 0.295 | -7.438 | **1.02E-13** |
| Organ specific autoimmune | 0.317 | 0.422 | -0.106 | 1.688 | 0.9 | 0.033 | 24.621 | -0.063 | 9.50E-01 |
| Renal Failure | -0.879 | -0.481 | -0.397 | 0.416 | 0.672 | 0.297 | 1.52 | -0.954 | 3.40E-01 |
| Stroke | -1.724 | -0.22 | -1.503 | 0.276 | 0.222 | 0.13 | 0.382 | -5.453 | **4.94E-08** |
| Systemic autoimmune | -0.362 | 0.069 | -0.43 | 2.423 | 0.65 | 0.006 | 75.146 | -0.178 | 8.59E-01 |
| T2DM | -1.533 | -0.789 | -0.745 | 1.31 | 0.475 | 0.036 | 6.189 | -0.568 | 5.70E-01 |
| VTE | -0.853 | -0.422 | -0.432 | 1.391 | 0.649 | 0.042 | 9.93 | -0.31 | 7.56E-01 |

Please refer to relevant abbreviations of table 1. Results with $p < 0.05$ are in bold.
Beta-post, coefficient from Cox regression in the post-vaccination period; beta-pre, coefficient from Cox regression in the pre-vaccination period; beta-diff, difference in coefficients in the post- and pre-vaccination periods. The PERR approach adjusts for (baseline) differences of the two groups prior to the introduction of vaccination, hence reducing unmeasured confounding.
1000 bootstraps were used to derive the standard error (boot-se) of beta-diff.

**Figure 3.**
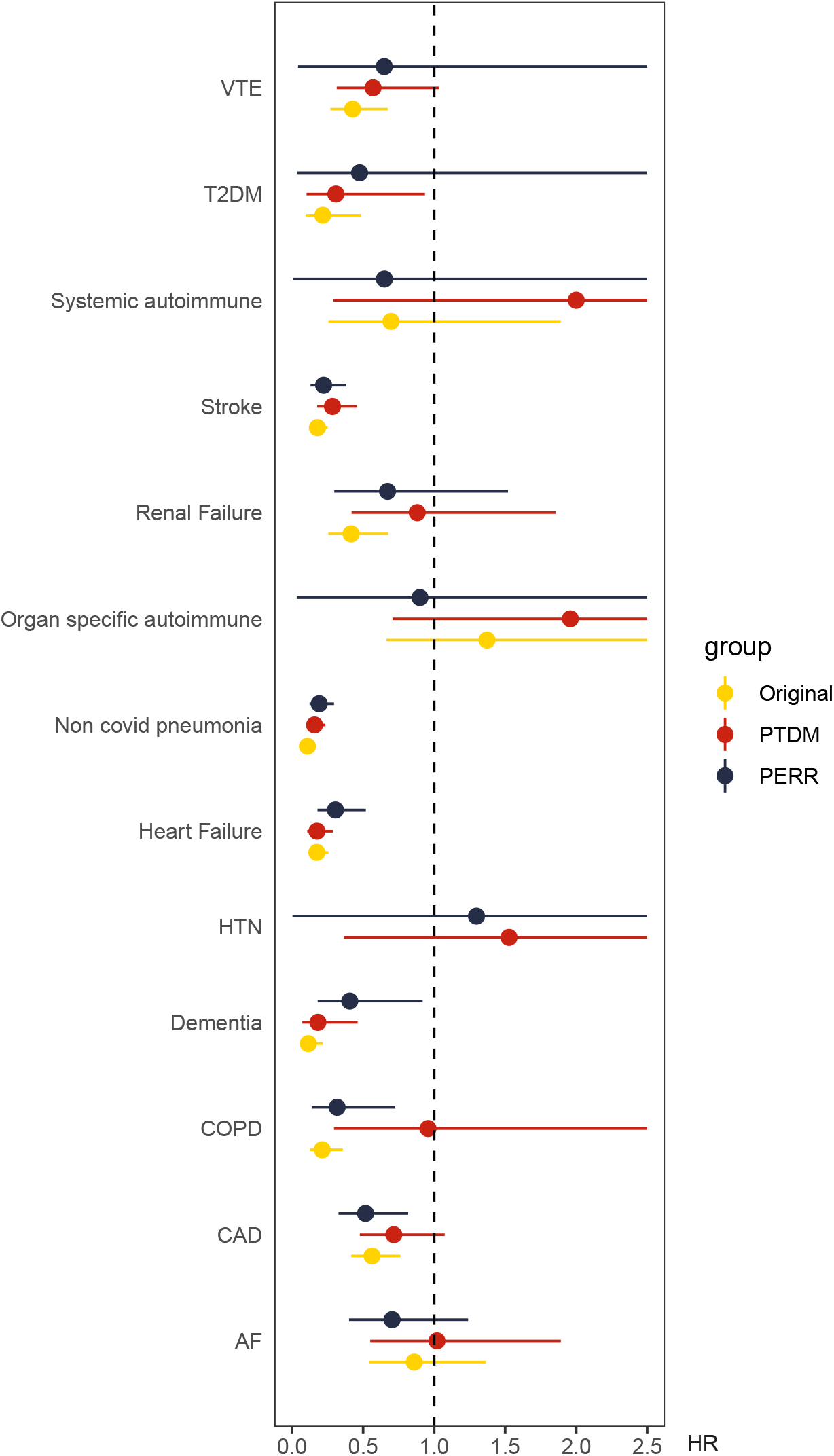
Association of (at least one dose of) COVID-19 vaccination with hospitalization from various diseases, based on the standard (original) approach, and adjustment by prior event rate ratio (PERR) or prescription time distribution matching (PTDM) approaches. The hazard ratio (HR) is shown on the x-axis. To preserve readability of the graph, the upper 95% confidence intervals of HR are truncated at 2.5.

Additionally, we performed PERR with two other one-year ‘prior’ periods(1-Jan-2019 to 7-Dec-2019 and 8-Dec-2019 to 7-Dec-2020 respectively); the results were largely similar. Full results are shown in Table_S6. Due to high computational cost, we performed PERR analysis only for the advanced model with covariate selection.

### Other sensitivity analyses

*PTDM adjustment* The main results with PTDM adjustment are presented in Table_3. Under PTDM adjustment(no PERR), reduced hazards of hospitalization were observed for dementia(HR=0.182,CI:0.072-0.461;p=3.21e-04), heart failure(HR=0.175,CI:0.107-0.286;p=3.88e-12), NCP(HR=0.158,CI:0.107-0.234;p=3.14e-20), stroke(HR=0.284,CI:0.177-0.456;p=1.92e-07) and T2DM(HR=0.308,CI:0.102-0.935;p=3.77e-02). As discussed earlier, the PTDM approach may tend to be conservative in this study which focused on hospitalization(instead of mortality) as the outcome. This purpose of the adjustment is to provide an extra sensitivity analysis to examine the robustness of findings.

**Table 3.** Association of (at least one dose of) COVID-19 vaccination with hospitalization from various disorders with PTDM (prescription time distribution matching) adjustment

| Outcome | Coef | Exp(coef)<br>[HR] | Se(coef) | Z | P | LCI | UCI | No of<br>Persons<br>unvac | Rate of<br>events<br>in<br>unvac<br>(per<br>100,000<br>person-days) | No of<br>Persons<br>vac | Rate of<br>events<br>in vac<br>(per<br>100,000<br>person-days) |
| --- | --- | --- | --- | --- | --- | --- | --- | --- | --- | --- | --- |
| AF | 0.02 | 1.02 | 0.315 | 0.064 | 9.49E-01 | 0.55 | 1.892 | 11 | 0.90 | 274 | 1.44 |
| CAD | -0.335 | 0.716 | 0.207 | -1.617 | 1.06E-01 | 0.477 | 1.074 | 27 | 2.22 | 443 | 2.33 |
| COPD | -0.044 | 0.957 | 0.601 | -0.073 | 9.42E-01 | 0.295 | 3.109 | 3 | 0.25 | 63 | 0.33 |
| Dementia | -1.702 | 0.182 | 0.473 | -3.598 | **3.21E-04** | 0.072 | 0.461 | 6 | 0.49 | 28 | 0.15 |
| Heart Failure | -1.745 | 0.175 | 0.251 | -6.942 | **3.88E-12** | 0.107 | 0.286 | 21 | 1.73 | 109 | 0.57 |
| HTN | 0.423 | 1.527 | 0.732 | 0.579 | 5.63E-01 | 0.364 | 6.409 | 2 | 0.16 | 60 | 0.32 |
| Non covid pneumonia | -1.846 | 0.158 | 0.2 | -9.214 | **3.14E-20** | 0.107 | 0.234 | 36 | 2.96 | 139 | 0.73 |
| Organ specific<br>autoimmune | 0.672 | 1.959 | 0.52 | 1.294 | 1.96E-01 | 0.707 | 5.426 | 4 | 0.33 | 125 | 0.66 |
| Renal Failure | -0.126 | 0.881 | 0.38 | -0.332 | 7.40E-01 | 0.419 | 1.856 | 10 | 0.82 | 112 | 0.59 |
| Stroke | -1.259 | 0.284 | 0.242 | -5.207 | **1.92E-07** | 0.177 | 0.456 | 21 | 1.73 | 173 | 0.91 |
| Systemic autoimmune | 0.775 | 2.17 | 1.025 | 0.756 | 4.50E-01 | 0.291 | 16.173 | 1 | 0.08 | 44 | 0.23 |
| T2DM | -1.176 | 0.308 | 0.566 | -2.078 | **3.77E-02** | 0.102 | 0.935 | 4 | 0.33 | 27 | 0.14 |
| VTE | -0.562 | 0.57 | 0.304 | -1.846 | 6.49E-02 | 0.314 | 1.035 | 13 | 1.07 | 135 | 0.71 |

Please refer to the legends of table 1.

*Different start-dates* We also repeated all our analyses(hospitalization as outcome) with the start-date defined as 14 days after(first) vaccination, as the vaccine may take longer to exert protective effects for infection. The results were largely similar to the above and are shown in Tables_S4-5. Significant findings were similar to the primary analyses.

*Subjects with no known infection* In another secondary analysis, we limited our analysis to subjects with no known history of COVID-19 infection all along. Here we present the results with PERR adjustment(prior period set to 1-Jan-2019 to 7-Dec-2020), which controls for residual confounding. We observed reduced hospitalization hazards from CAD(HR=0.663,CI:0.447-0.982,p=4.04e-02), COPD(HR=0.465,CI:0.246-0.877,p=1.80e-02), heart failure(HR=0.502,CI:0.324-0.778,p=2.05e-03), NCP(HR=0.290,CI:0.201-0.420,p=5.24e-11) and stroke(HR=0.321,CI:0.203-0.508,p=1.20e-06)(Table_S4-5). There was no significant association of vaccination with dementia. The HR estimates were higher(but all HR<1) when restricted to subjects with no known infection, indicating weaker protective associations.

There was no evidence of violation of the proportional hazards(PH) assumption for the majority of Cox regression results(all p>0.05,Table_S9).

### Different doses and types of COVID-19 vaccine

The pattern of associations were similar for single-dose and at least one dose of vaccination(Table_S10, Figure_S9), when compared to no vaccination. Although the group with two doses was limited in size, we still observed reduced hazards of hospitalization for most outcomes(including NCP/stroke/HF/COPD/CAD/AF/RF/dementia/VTE). For analysis restricted to new-onset or relapse/recurrent cases, results were largely consistent(Table_S10).

We did not find statistically significant differences between two-dose and one-dose vaccination generally(Table_S10, Fig_S9). However, for AF, significant reduction in hospitalizations was only observed with two doses (HR=0.547,CI:0.312-0.958), when compared to the unvaccinated. Besides, we observed that two-dose vaccination was associated with modestly higher risks of hospitalization from VTE(HR=1.539,CI:1.022-2.320;p=3.91e-02), compared to one-dose only(Table_S10/Fig_S9). Nevertheless, both two-dose and one-dose vaccination were actually associated with reduced VTE hospitalizations(two-dose_HR=0.546[0.306-0.977]; one-dose_HR=0.400[0.245-0.651]) compared to the unvaccinated.

Regarding the different types of vaccines, the patterns of hospitalization risk reduction were similar for BioNTech and AstraZeneca vaccines(Table_S11;Fig_S10-11). When comparisons were made between the two vaccines, we found a marginally significant result for VTE for BNT162b2 vaccine(HR=1.625,CI:1.000-2.642;p=4.99e-02)(Table_S11;Fig_S10-11).

## Discussions

### Overview of main findings

To our knowledge, this is one of the most comprehensive studies to date on the association between COVID-19 vaccination and hospitalization from cardiovascular and other diseases, and among the first to report possible protective associations. Overall, we observed that COVID-19 vaccinations(at least one-dose) was associated with lower risks of hospitalizations from several diseases in short-term follow-up. In particular, reduced hospitalizations for stroke, heart failure, NCP and dementia were consistently observed across different analyses, including after PERR adjustment. Reduced hazards of hospitalization was also observed for CAD and COPD. We also observed decreased hospitalizations risks for VTE, renal failure and T2DM in our analyses, which may be worthy of further investigations, although the associations were non-significant after PERR adjustment. In general, there is no evidence that vaccination was associated with increased hospitalization from the disorders studied, providing support for the safety of COVID-19 vaccines.

### Interpretation of findings

There is unequivocal evidence that both the BioNTech and Oxford-AstraZeneca vaccines were effective in reducing risks of COVID-19 infection and severe disease, and partially vaccinated individuals are also protected, especially against severe disease^26,27^. There is mounting evidence that COVID-19 is associated numerous sequelae^28,29^. For instance, studies showed that COVID-19 was associated with elevated risks of stroke^30,31^, VTE^16^, heart failure^32^ and other cardiac disorders^33^, renal dysfunction^34^, bacterial pneumonia^35^ and mortality associated with dementia^18^. COVID-19 is also associated with higher cardiovascular and all-cause mortality, up to 6 months after the infection^29^. Taken together, COVID-19 vaccines may partially protect against hospitalization from some of these disorders, likely via reducing infection and severe disease. An analogy may be drawn with influenza vaccination. As described earlier, numerous studies showed that flu vaccination was associated with reduced cardiovascular risks/mortality, despite that flu vaccines generally have lower efficacy and risks of severe disease/complications from flu are lower than those from COVID-19.

A recent nationwide study^36^ examined the safety and potential adverse effects of the BNT162b2 vaccine with a follow-up period of ∼42 days. Considering adverse effects, the largest effect size was observed for myocarditis(HR=3.24,CI:1.55–12.44). We did not cover myocarditis here due to low incidence of the condition. Although not the primary focus of the study, the authors also reported significantly reduced incidence of several conditions, including acute kidney injury, anemia, intracranial hemorrhage and other thrombosis^36^. We also observed reduced hazards of renal failure, VTE and stroke hospitalizations after vaccination in this study, corroborating with previous findings. Another study using an adverse event database(Vigbase) reported possible link of vaccination with myocardial infarction, cardiac arrest and hypertensive emergencies and some arrthymias^37^; however, these findings were based on disproportionality analysis which did not control for any confounders and based on spontaneously reported events only; higher tendency to report events after COVID-19 vaccines compared to other vaccines may also bias the results.

Interestingly, we observed that the protective associations for several diseases(including CAD, COPD, heart failure, NCP and stroke) remained after restriction to subjects with no known history of infection all along, although the effect sizes were attenuated. One possible explanation is that subjects with mild/moderate infections may not get tested(or reported), but such infections can still be associated with elevated risks of complications compared to the non-infected^29^, especially in older adults. COVID-19 vaccination also protects against non-severe infections^27,38^ and hence may also protect against the corresponding complications. An alternative possibility is that the vaccine may provide beneficial effects via other mechanisms beyond protection from COVID-19. For flu vaccine, pre-clinical studies showed that it may stabilize atherosclerotic plaques and increase nitric oxide production^12^. However, the exact mechanisms require further investigations. Of note, some studies have shown that vaccines may confer non-specific protection against diseases not originally targeted by the vaccine. For example, several studies reported benefits of flu vaccines against infection or severe infection from COVID-19^39-41^, and other vaccines e.g. MMR or BCG may also protect against COVID-19^42-44^ through ‘training’ of innate immunity. One may hypothesize that COVID-19 vaccines may also confer some non-specific protective effects against other infections such as other types of pneumonia.

We also performed further exploratory analyses comparing different doses and types of vaccines. We did not find significant differences between one or two doses of vaccination, suggesting even one dose of vaccination (compared to none) may be useful in reducing hospitalizations from other disorders, possibly via reduction of severe infections. Nevertheless, lack of significant differences can be due to inadequate power for smaller differences. Also, other variants of concern(e.g. Delta/Omicron) were not studied, and immune escape from some variants may necessitate further doses to improve efficacy. Regarding vaccine types, again we did not find significant differences between AstraZeneca and BioNtech vaccines generally. Similar effects for preventing hospitalization/mortality were observed for both vaccines in early studies^26^. Nevertheless, the power is limited for small differences and whether there are differences in the longer term or for other variants remains unknown.

### Strengths and limitations

This study is based on a large sample with detailed phenotypic information and health records. We conducted a comprehensive analysis covering a wide range of cardiovascular and other relevant diseases. We also conducted analyses under different statistical models to evaluate if the findings are robust to different modeling strategies. For example, the PERR method adjusts for HR before introduction of vaccines to minimize residual confounding.

There are several limitations. Firstly, this is a real-world observational study without randomization. As such, residual confounding cannot be excluded, and causality cannot be confirmed. Possible confounders may include, for instance, differences in health-consciousness or health-seeking behaviors between the vaccinated and non-vaccinated. We have employed the PERR approach to reduce unmeasured confounding, and it is reassuring that a number of associations remained significant after the adjustment.

Another limitation is that the number of events is relatively small in view of short follow-ups. This study is not intended to cover all possible medical conditions; vaccination can be possibly associated with rare adverse events(e.g. myocarditis) not covered here. Also, longer-term effects of vaccinations remains to be addressed. A related limitation is that relatively few events were observed in fully vaccinated individuals(and the time available for follow-up is much shorter), hence we focused on the effects of at least one dose of vaccine. We note that the status of vaccination changes rapidly, for example booster doses are already widely implemented. Nonetheless, we believe that the main conclusion that COVID-19 vaccination may reduce risks of hospitalization from other disorders is unlikely to be changed, as such protective benefit is likely to extend to boosters. Also, from an international perspective, only ∼24.5% of people in low-income countries have received at least one dose (https://ourworldindata.org/covid-vaccinations, accessed 25-Nov-2022). Another point to note is that vaccine effectiveness for infection may wane over time^45,46^; it is unknown whether the effect in preventing the studied diseases will also attenuate over time. However, current evidence suggests that effectiveness against severe infections remains at relatively satisfactory levels^45,46^. The emergence of viral variants may affect vaccine effectiveness^47^ but variant information is currently unavailable. Nevertheless, if the vaccine is still effective against severe infections, protection from hospitalization from other disorders will likely hold. Finally, the UKBB may not be representative of the entire UK population^48^. Also, generalizability of the findings to other types of COVID-19 vaccines and other populations(e.g. different age-groups/ethnicities) remains to be studied.

### Implications

If our findings are replicated, they may have several clinical and public health implications. Importantly, this study provides further support to the safety and benefits of COVID-19 vaccination. We observed that COVID-19 vaccines are possibly associated with lower risks of hospitalization from several cardiovascular, respiratory and other diseases; this may provide important further incentives for vaccination, especially those worried about underlying medical conditions. In addition, the current findings suggest additional benefits for patients with certain comorbidities, which may have implications for prioritization of vaccination in resource-limited settings. For example, only ∼17.4% of people in low-income countries have received at least one dose of COVID-19 vaccination^49^(accessed 30-Jun-2022).

## Conclusions

In conclusion, the current study suggests that COVID-19 vaccination may protect against hospitalization from several diseases, such as stroke, heart failure, CAD, NCP, COPD and dementia. Taken together, this study provides further support to the safety and benefits of COVID-19 vaccination, and such benefits may be beyond reduction of infection risk or disease severity per se. As an observational study, causal relationship cannot be concluded and further studies are required to verify the findings.

## Data Availability

Qualified researchers may apply for access to the UK Biobank, which contains individual-level data. Summary results are presented in main and supplementary tables.

## Acknowledgements

This work was supported partially by a National Natural Science Foundation China(NSFC) grant(81971706) and the Lo Kwee Seong Biomedical Research Fund from The Chinese University of Hong Kong and the KIZ-CUHK Joint Laboratory of Bioresources and Molecular Research of Common Diseases, Kunming Institute of Zoology and The Chinese University of Hong Kong, China. We thank Prof. Pak Sham for support on data access.

## Authors’ contributions

Conception and design: HCS(lead), with input from YX and YF. Study supervision: HCS. Funding acquisition: HCS. Methodology: HCS(lead), YX. Data curation: YX(lead), YF, JQ. Data analysis: YX(lead), YF, JQ, RZ. Data interpretation: HCS, YX, YF. Preparation of first draft of manuscript: HCS(lead), YX, with input from YF and JQ.

## Data availability

All data used in the current analysis is available from the UKBB upon application.

## Supplementary Information

All supplementary Tables, Figures and notes are available at the journal’s website and at https://drive.google.com/drive/folders/1177cQAt2SMUXd136MTGGsqCVJ9gBLi8t?usp=sharing

